# Sociodemographic factors associated with knowledge of common eye diseases: a cross-sectional study in Afghanistan

**DOI:** 10.1101/2022.09.19.22280069

**Authors:** Farooq Hosaini, Bijaya Kumar Padhi, Parimala Mohanty, Nosaibah Razaqi, Mehrab Neyazi, Elhama Noorzad, Adina Rahmani, Marjan Yousufi, Habibah Afzali, Aritro Bhattacharyya, Ahmad Neyazi

## Abstract

**Background:** Eye diseases are a major global health concern but are often neglected. This study aims to assess the sociodemographic determinants of knowledge on common eye diseases among the adult population in Afghanistan.

**Methods:** A cross-sectional survey was designed and conducted between August and October, 2021, including 509 adult population in the Herat province of Afghanistan. Participants’ sociodemographics and knowledge of common eye diseases were recorded through an in-person interview. A multivariable logistic regression model was used to investigate factors associated with understanding common eye diseases among study participants.

**Result:** The mean age of the study participant was 34.96 years, with males making up nearly 53.4% of the total sample. Of the 509 respondents, 76.8% of them did not have enough information on common eye diseases; 79% of the participants did not know the definition of glaucoma disease. Only 47.3% of the participants of this study knew the meaning of cataract disease. The adjusted odds ratios (AOR) revealed that participants over 35 years of age had significantly higher odds of knowledge of eye disease [AOR: 3.629; 95%CI:1.770, 7.442] compared to the relatively younger age group of 18-34 years. A significant association was found between awareness about eye diseases and higher levels of education. Participants receiving higher education were found to have higher odds of being aware of the knowledge of eye disease.

**Conclusions:** Results showed low awareness about common eye disease among the younger adult population. The study, therefore, consolidates the need for promoting health literacy regarding common eye diseases in Herat province of Afghanistan.

## Introduction

Visual impairment, and even blindness are significant burdens to the productivity of a community, cutting off their social life and drying up their financial resources. The burden of visual impairment leading to loss of proper vision is a grim reality in many populations around the globe, more so in lower-income countries. Common ocular diseases like Glaucoma, cataracts, and diabetic retinopathy are cited globally as the leading causes of blindness. As stated by WHO, around 285 million people have visual impairment worldwide, 246 million have moderate or intense visual impairment, and 39 million are blind [1–3]. Of these, 19 million are children under 14 years old [4]. Common eye disorders include refractive errors, cataracts, age-related macular degeneration, Glaucoma, and trachoma [5–7]. Uncontrolled refractive errors (43%) are the principal cause of vision loss and are trailed by cataracts (33%), diabetic retinopathy, Glaucoma, and age-related macular degradation [8].

Opportunely, visual impairment in nearly 80% of cases is avoidable and curable [12]. Knowledge and awareness of common eye disorders assist people in asking for an ophthalmologist at the opportune time. Early diagnosis leads to effective therapy and prevents visual impairment [13–15]. Lack of proper, specified, and organized health awareness, added with the delay in early diagnosis and prevention, is often found to contribute largely to the prevalence of these visual complications. The impact of blindness and visual impairment is not only on the quality of life of human beings but also has a negative impact on their educational and career opportunities. Therefore, raising awareness and knowledge of common eye diseases encourages early diagnosis and reduces the burden of visual impairment in the long run [16].

According to studies, a strong association between socioeconomics and prevalence and causes of vision loss and blindness has attracted the attention of eye specialists and public eye care planners (9,10). Socioeconomic factors describe 69.4% of worldwide alteration in moderate to intense vision impairment and 76.3% of global alteration in the prevalence of sightlessness (11). The variant outcome has been worldwide awareness of common eye disorders (2,15,17,18). In developed countries like Canada, eye disease awareness statistics were stated at 69% for cataracts and 41% for Glaucoma. Whereas awareness about common eye disease in the civic population in India was low, for example, in a study from southern India, most patients (90%) with Glaucoma were unaware of the condition and its complication. Another study in India reported a poor awareness of Glaucoma (3.2%) and diabetic retinopathy (27%) in the study population (19). Some countries like Saudi Arabia and the United States have focused on studies related to age-related eye disease or specific eye problems, such as dry eye (20,21). However, in Afghanistan, prior studies have mostly focused on eye injury or blindness, and the body of evidence regarding the common eye disease in Afghanistan contains considerable gaps (22–24). According to the available research, knowledge and awareness of eye diseases in Afghanistan are scant and limited. In one study conducted among British Armed Forces in Afghanistan, the rate of ocular injuries was recorded (22). In another study, Afghanistan military hospital reported Combat Ocular Trauma injuries (23). Further blindness, vision impairment, and cataract prevalence were studied in Kabul (24).

Currently, there is insufficient data regarding the adult population’s awareness of common eye diseases in Afghanistan among a special category of people living. This study aimed to assess the overall understanding of the concerned population on common eye diseases and the association of the same with existing sociodemographic features.

## Methods

### Study design, sample, and settings

The cross-sectional study on the public’s awareness of common eye diseases was conducted among 509 adult population (18 years or above) of Herat province in Afghanistan. The sample size (N=509) was calculated using OpenEpi Version 3.01 with a hypothesized 50% ± 5% prevalence of knowledge on common eye diseases with a 95% confidence level. The design effect was kept one. The minimal sample size, which came on estimation at a 95% confidence level, was 384. To account for non-responses, we recruited 509 participants aged 18 and above for data collection.

### Data collection

A print-based Dari language questionnaire was used in this study. Prior initiating the data collection process, the drafted questionnaire was distributed to twenty random people to review and check for the comprehensibility of the questions. It resulted in a minor revision of the questionnaire to make its included questions understandable to common people. The final questionnaire consisted of two sections: the sociodemographic section, and the awareness-related items section.

Sociodemographic information included questions on age, gender (male, female), marital status (single, married, widow/divorced), location (urban, rural), education (illiterate, primary school, secondary school, high school, university), and occupation (occupied, searching for a job, housewife, other).

The awareness-related items section included four open-ended questions to assess the participant’s basic knowledge of Glaucoma, cataract, diabetic retinopathy, and dry eye. Each of the questions had three sections and scored from 0 to 3. Knowing about each of the diseases is related to eye score 1. Knowing risk factors of the disease were scored 1. Knowing the disease’s cause for blindness, preventability, curability, and reversibility were scored 1 as well. Each of the participants could score from 0 to 12. Participants with a knowledge score of 0 to 6 were considered “not aware”, and participants with a knowledge score of 7 or above were considered “aware”.

Data were collected from 15 districts of Herat city of Afghanistan. The convenience sampling technique was used to reach the participant. Each of the participants was interviewed by the data collectors. A total of 509 adults aged 18 years or above participated in this study.

People who agreed with the terms and conditions of participation in the study were selected with the knowledge that they can select not to continue at any time during the progress. Male or females living in Herat province who agreed to provide an verbal agreement and aged above 18 years old were included in this study.

### Statistical Analysis

Data processing and analyzing the data were done in IBM SPSS version 26. We used the frequency option to obtain the numbers and percentages for the sociodemographic variables and factors connected to prevalent eye disorders. Similarly, to evaluate the relationship between different variables, chi-square tests were used. Logistic regression models were used to compute the odds ratio for establishing an association between sociodemographic factors and knowledge of eye diseases among study participants. A *p*-value of less than 0.05 was considered significant in the present study.

### Ethical Consideration

The Afghanistan Center for Epidemiological Studies’ Ethical Committee approved the study protocol (reference number #21.0020; August 1, 2021). The entire process and objective of the study were explained to the participants during the initial contact. All procedures were carried out in compliance with the applicable ethical standards and guidelines.

## Results

This study included 509 adult participants, ranging in age from 18 to 78 years. The study participants had a mean age of 34.96 years. Male participants made up 53.4% of the total participants. Nearly two third of the people (65.0%) who participated in the study were in married relationships. Four out of five participants were living in urban districts of Herat city. More than one-third of the participants were illiterate (37.5%). One out of three participants was a housewife (37.7%). [**Table 1**]

**Table 1:** Characteristics of study participants (N=509)

| Characteristic | Category | N | (%) |
| --- | --- | --- | --- |
| Age group | 18 – 34 years | 276 | 54.2 |
|  | ≥ 35 years | 233 | 45.8 |
| Gender | Male | 272 | 53.4 |
|  | Female | 237 | 46.6 |
| Marital status | Single | 159 | 31.2 |
|  | Married | 331 | 65.0 |
|  | Widow/divorced | 19 | 3.7 |
| Location | Urban | 402 | 79.0 |
|  | Rural | 107 | 21.0 |
| Education | Illiterate | 191 | 37.5 |
|  | Primary school | 49 | 9.6 |
|  | Secondary school | 41 | 8.1 |
|  | High school | 89 | 17.5 |
|  | University | 139 | 27.3 |
| Occupation | Occupied | 194 | 38.1 |
|  | Searching for job | 65 | 12.8 |
|  | Housewife | 192 | 37.7 |
|  | Other | 58 | 11.4 |
| **Total** |  | **509** | **100.0** |

Table 2 shows study participants’ knowledge on eye diseases in Afghanistan. Generally, almost three out of four participants did not have enough information on common eye diseases (76.8%). Nearly four out of five participants did not know the definition of glaucoma disease (79.0%). Less than half of the participants of this study knew the meaning of cataract disease (47.3%). Almost one out of seven participants knew the true definition of diabetic retinopathy disease (15.5%). About half of the participants were unaware of the risk factors of Glaucoma (47.9%) and dry eye diseases (50.3%). [**Table 2**]

**Table 2:** Study Participants Knowledge on eye diseases (N=509)

| Knowledge on: | Aware (%) | Not aware (%) |
| --- | --- | --- |
| Glaucoma | 107 (21.0) | 402 (79.0) |
| Cataract | 241 (47.3) | 268 (52.7) |
| Diabetic retinopathy | 79 (15.5) | 430 (84.5) |
| Dry eye | 121 (23.8) | 388 (76.2) |
| Glaucoma risk factors | 265 (52.1) | 244 (47.9) |
| Cataract risk factors | 307 (60.3) | 202 (39.7) |
| Diabetic retinopathy risk factors | 258 (50.7) | 251 (49.3) |
| Dry eye risk factors | 253 (49.7) | 256 (50.3) |
| Causes blindness | 112 (22.0) | 397 (78.0) |
| Preventable | 54 (10.6) | 455 (89.4) |
| Curable | 240 (47.2) | 269 (52.8) |
| Reversible | 50 (9.8) | 459 (90.2) |
| **Total** | **118 (23.2)** | **391 (76.8)** |

**Table 3** represents the multivariable regression estimates for participants’ knowledge of eye disease by their background sociodemographic variables. The first model represents the unadjusted odds ratio, and the second model is described the adjusted odds ratio. The AOR revealed that participants over 35 years of age had significantly higher odds of knowledge of eye disease [AOR: 3.62; 95%CI:1.770, 7.442]. Even participants’ education had significantly higher odds of knowing the eye disease, i.e., secondary school [AOR: 3.43; 95%CI: 1.35, 8.698], high school: [AOR: 2.81; 95%CI: 1.160, 6.825], and university: [AOR: 10.25; 95%CI: 4.234, 24.840] respectively.

**Table 3:**
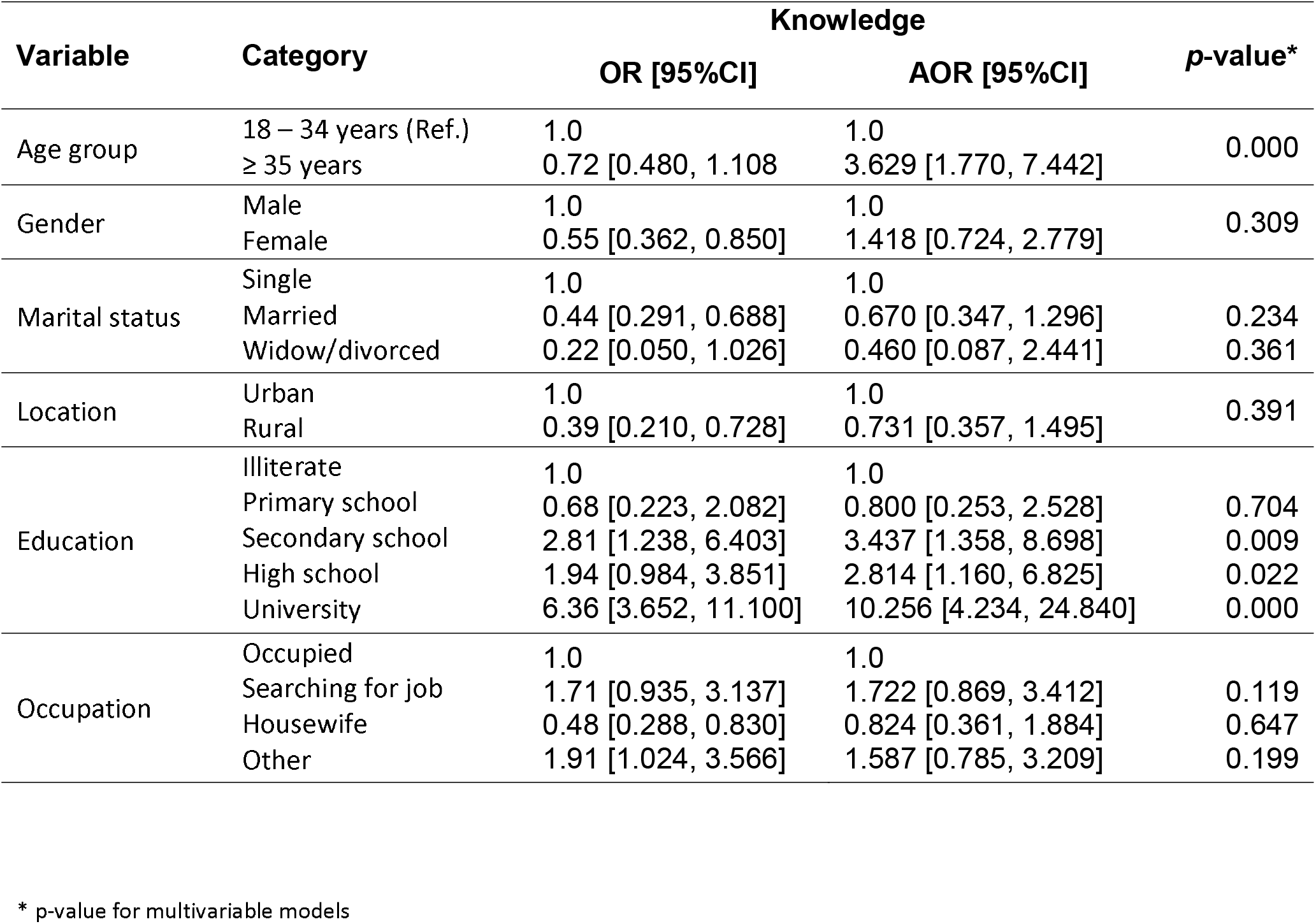
Association between participants’ knowledge on eye disease and their selected socio-demographic variables (N=509)

## Discussion

Our research is the first of its kind to determine awareness and knowledge concerning common eye diseases among the adult population of Herat city, Afghanistan. The key takeaway from our research is that people living in Herat city have a limited understanding of prevalent common eye diseases. Around three-fourths of the survey participants didn’t have enough information on common eye diseases. Nearly eighty percent of the participants did not understand the meaning of the term “glaucoma disease.” Only 15% of the sample correctly defined diabetic retinopathy as a condition. Nearly half of the participants did not know the causes of Glaucoma or dry eye syndrome. However, slightly more than half of the people in this survey correctly defined cataract illness (51.3%). Those with less education and younger in age were particularly vulnerable to this shortcoming. The findings are helpful in far more than one way. There are mainly two takeaways from this research. Firstly, it shows that the general public has a very inadequate understanding of the most common eye disorders. Secondly, this deficit is exacerbated in very young populations and low-literate comparable to other developing nations such as Bangladesh and Pakistan (7,8).

Demonstrating the frequency of different eye diseases in our population was outside the scope of this investigation. Still, evidence suggests that raising awareness and education about the importance of annual eye exams can help minimize ocular diseases and the resulting financial costs (13).

Our research findings were different from those of a similar study conducted in Pakistan. In our research, Glaucoma had the lowest awareness (79.0%) and the highest understanding (60.3%) of the cataract risk factor, whereas, in the study by Zhao et al., the age-related macular degradation had the lowest awareness (31.4%) and the highest level of understanding of the fact that blindness can be prevented (68.1%) (8). Further, in contrast to the above-said study, only 10.6% of the research participants in our study were aware that blindness can be prevented.

Comparable to our study, Islam et al. found the lowest awareness for diabetic retinopathy and the highest awareness for cataracts (7). However, acceptable awareness in both studies was low and was mostly linked with the education level of the participants (7,8). In this study, 47.3% and 50.7% of participants were aware of cataracts and diabetic retinopathy, respectively. These findings differ from those of the study by Marzieh et al., which revealed that 82.9% of participants knew about cataracts and 86.2% knew about diabetic retinopathy.

In this study, awareness of cataracts and diabetic retinopathy were at 47.3 and 50.7%, respectively. While Marzieh et al. found that 82.9% of participants were aware of cataracts and 86.2% were aware of diabetic retinopathy, these results contradict our findings (10). The cataract findings of our study also differ from those of Lau et al., who found that 90% of respondents were aware of the condition (25).

In terms of the association of participants’ knowledge of eye disease with the socio-demographics, in our study population over the age of 35 years or more was 3.62 times (AOR = 3.62 95%CI = 1.770, 7.442) more aware as compared the population in the age group 18–34 years. These results were consistent with Dandona’s earlier findings that the probabilities of being aware rise with age, showing that people in the 60-69 age group were 3.25 times (AOR = 3.25 95%CI = 2.10-5.0) more knowledgeable than people in the 16-29 age group (13). However, another study found that the elderly were 1.21 times (AOR = 1.21 95% CI = 0.56, 2.61) more likely to be aware of trachoma than the young adults (age 18-30) (8). Further, our study’s results corroborate a significant association between greater awareness and increasing education levels, as demonstrated by previous research (7,7,8,13,25). Other researches reveal distinctions in male and female awareness levels (10,14). In the current study, however, there were no significant gender differences in eye disease knowledge and awareness.

Interestingly, a study conducted in Bangladesh found no statistically significant difference in the levels of knowledge held by men and women, which stands similar to our study (7). While it is interesting to draw parallels between the findings of the other similar studies, which included either only rural or a mix of both rural and urban residents, it cannot be compared with our study, where a majority of our population lives in urban areas, and their average socioeconomic and educational levels are different. However, it is seen that populations with lower levels of education, younger age, and low socioeconomic status were more likely to lack knowledge about eye diseases in both emerging and developed nations (7,14,26).

Findings from this study provide credence to the argument that a greater effort should be made to improve health literacy about eye disease among people of all socioeconomic backgrounds, but notably among the younger population and those with lower levels of education. This is particularly crucial in nations such as Afghanistan, where there is a shortage of health care professionals (27).

Our study has some limitations; firstly, study participants were recruited using a convenience sampling approach. Further, it was beyond the scope of this study to determine the prevalence of eye illness. Still, we believe that raising awareness of the importance of routine eye exams will help reduce both visual impairment and the overall expense of eye care. Finally, we were unable to determine the interviewer’s dependability. Our time and resources did not permit two interviewers to examine inter-rater reliability. However, data collection was performed by skilled and trained enumerators and was double-checked by a senior researcher for completeness and accuracy.

Despite the above limitations, this is the first survey from Afghanistan of its kind; the current study provides a clear picture of the awareness of prevalent eye disorders among the general population in Afghanistan. The study used in-person interviews conducted by competent researchers to ensure the accuracy and reliability of its findings.

## Conclusion

Results from the study demonstrated an overall low prevalence of awareness and knowledge of common eye diseases among the adult population of Herat province, Afghanistan. Young adults were found to be relatively more unexposed to information on eye diseases compared to the older population. In contrast, the more educated were found to have higher odds of being aware than the less educated. The findings strongly suggest improvements in health literacy and public interventions, specifically in young people with lower education.

## Data Availability

All data produced in the present study are available upon reasonable request to the authors

## Author contributions

All authors contributed substantially to the conception and design, data collection and entry; data analysis and interpretation; drafting of the article. All authors approved the manuscript for publication.

## Ethical Consideration

The Afghanistan Center for Epidemiological Studies’ Ethical Committee gave their clearance for this study (reference number #21.0020; August 1, 2021). All procedures were carried out in compliance with the applicable ethical standards and guidelines. A detailed description of the study, benefits, confidentiality was explained during the initial contact with the participants prior to their participation. Verbal agreement was recruited from all the participants involved in this research.

## Conflict of interest

The authors declare no conflict of interest.

## Funding

This research received no external funding.

## Acknowledgment

We would like to express our sincere gratitude to all the participants in this study.

## Notes

### Competing Interest Statement

The authors have declared no competing interest.

### Funding Statement

This study did not receive any funding

